# Differential COVID-19 case positivity in New York City neighborhoods: socioeconomic factors and mobility

**DOI:** 10.1101/2020.07.01.20144188

**Authors:** Matthew R. Lamb, Sasikiran Kandula, Jeffrey Shaman

**Affiliations:** Department of Epidemiology, Mailman School of Public Health, Columbia University, New York, New York; ICAP, Mailman School of Public Health, Columbia University, New York, New York; Department of Environmental Health Sciences, Mailman School of Public Health, Columbia University, New York, New York

**Keywords:** COVID-19, socioeconomic status, mobility, generalized estimating equations, New York City

## Abstract

New York City has been one of the hotspots of the COVID-19 pandemic and during the first two months of the outbreak considerable variability in case positivity was observed across the city’s ZIP codes. In this study, we examined: a) the extent to which the variability in ZIP code level cases can be explained by aggregate markers of socioeconomic status and daily change in mobility; and b) the extent to which daily change in mobility independently predicts case positivity.

Our analysis indicates that the markers considered together explained 56% of the variability in case positivity through April 1 and their explanatory power decreased to 18% by April 30. Our analysis also indicates that changes in mobility during this time period are not likely to be acting as a mediator of the relationship between ZIP-level SES and case positivity. During the middle of April, increases in mobility were independently associated with decreased case positivity. Together, these findings present evidence that heterogeneity in COVID-19 case positivity during the New York City spring outbreak was largely driven by residents’ socioeconomic status.

---

New York City (NYC) was severely impacted by the global COVID-19 pandemic, with a reported 164,505 cases, 42,417 hospitalizations and 13,000 laboratory-confirmed deaths through April 30, 2020(1). These represented 16.4% (5.3%) of cases and 24.8% (5.9%) of deaths nationally (globally)(2). The COVID-19 case positivity (the fraction of viral diagnostic tests that were positive) has been heterogeneous across the city’s neighborhoods (3). There is also considerable variability among different ZIP codes within the boroughs (4).

Potentially opposing mechanisms may help understand differentials in the case positivity proportion between wealthier and less wealthy ZIP codes: access to the COVID-19 diagnostic tests themselves and the underlying true (but imperfectly measured) COVID-19 prevalence by ZIP code. First, individuals living in wealthier ZIP codes may have found it easier to circumvent the restrictive initial testing guidelines on eligibility for a COVID-19 diagnostic test, resulting in a lower proportion receiving the test actually being COVID-19 positive. Conversely, individuals living in less wealthy ZIP codes may have been less able to receive tests unless clinically sick due to a lower proportion having a primary care physician and therefore reliant on emergency care for clinical consultation, suggesting that individuals living in poorer neighborhoods who eventually receive tests are more likely to be COVID-19 positive. Second, evidence strongly suggests that the actual prevalence of COVID-19 is substantially higher among Black individuals and those of lower socioeconomic status (5, 6). There are many potential explanations for these disparities. Individuals living in wealthier neighborhoods may have greater ability to reduce personal exposure by abiding by social distancing guidelines per the *New York State on PAUSE* directive (7), transition to work-at-home, limit visits to stores by having essentials delivered, reduce usage of public transportation, and may be more able to shelter outside of NYC.

This analysis utilizes routinely-reported public data from the New York City Department of Health and Mental Hygiene (NYC DOHMH), coupled with anonymized cell phone data assessing frequency of visits to businesses and U.S. Census data to examine (1) the extent to which heterogeneity in the COVID-19 daily case-positivity proportion by ZIP code can be explained with aggregate markers of socioeconomic status (SES) along with markers of daily change in mobility; and (2) the extent to which daily change in mobility independently predicts COVID-19 daily case-positivity proportions.

## METHODS

In this ecologic study, our study population consists of aggregate data collected among residents in 177 ZIP codes of New York City covering all five boroughs. Our primary outcome of interest is the proportion of COVID-19 tests found positive in each ZIP code. These data were extracted from a versioned public repository updated daily by the NYC DOHMH (8) with the first release date on April 1, 2020. Positivity on a given day is calculated as the fraction of new tests during the last three days found positive. The use of a moving window rather than raw counts on a single day smooths out fluctuations due to reporting constraints, especially around weekends when fewer tests are sought, administered and/or reported than on weekdays.

ZIP code-level characteristics used as explanatory variables in this study were extracted from the U.S. Census and the American Community Survey 2016 (ACS; codes in parentheses) and include:

- Proportion of the 18-64 year old population that is uninsured (B27010),
- Median household income (in 2016 dollars, B19013)
- Proportion of population that self-identified their race as white (B02001)
- Proportion of population living in households with more than three inhabitants (B11016)
- Proportion of population using public transportation to commute to work that includes bus travel (B08301)
- Proportion of population that is elderly (65+ years of age) (B01001)

Anonymized location data from cell phone visits to businesses within a ZIP code were obtained via SafeGraph (9, 10). Across NYC, SafeGraph provides the location (to the resolution of U.S. Census block group) of one of approximately 75,000 points of interest (POIs) and the number of daily visits to each POI as tracked by mobile phones. POIs are defined by SafeGraph as “a specific physical location which someone might find interesting” and includes businesses, workplaces, educational institutions, and transit centers. These data were chosen in part to provide a comprehensive assessment of visitation patterns in a given ZIP code and for comparability with mobility estimates released by CDC (11). For this analysis, we aggregated the total number of visits to all POIs in a ZIP code on a given day and divided by the number of POIs in that ZIP code to estimate total visits per business per day (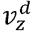, visits per in ZIP code *z* during day *d*). As a measure of baseline or reference mobility 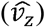, we calculated median daily visits per during pre-pandemic period, defined as a 6 month period from September 2019 to February 2020. Our exposure of interest, referred to henceforth as *mobility*, is the proportional change from pre-pandemic mobility for each ZIP code, operationalized as the proportion of baseline mobility experienced in a given day of post-pandemic response, 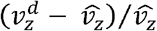,. To account for the estimated incubation period of COVID-19(12) we lagged the mobility variable by 7 days. An examination of mobility trends showed considerable variability in baseline mobility across ZIP codes and the rate at which mobility changed over time. To examine potential differential impacts across time, we report our results at four time points (April 1^st^, 10^th^, 20^th^, and 30^th^) with the April 1^st^ time period reported as cumulative cases for the month of March.

### Statistical analyses

Our first goal was to build a model estimating the proportion of total variability in percent positivity by ZIP code that could be explained by aggregate measures of SES. We considered ZIP code-level measures of SES to be time-invariant across the time period of the study (April 1 - April 30). By the end of April a consistent downward trend in daily cases was evident and testing was widely available. Therefore we limited the study period to April. As percent positivity changed daily during the study period, multivariable linear regression with generalized estimating equations with an autoregressive correlation structure were used to account for within-ZIP clustering, assuming that outcome measurements within each ZIP code were more highly correlated with each other than across ZIP codes, and that the strength of this correlation was dependent on the time separation between measures. ZIP codes were assumed to be independent of one another. Standard errors reported are robust estimates. First, ZIP code-level measures were considered in univariate analyses and ranked according to the percentage of variability explained (using R^2^). A multivariable prediction model was then constructed by sequentially adding variables in order from the highest to the lowest R^2^ from univariate analysis, until the adjusted R^2^ changed by less than 5% or all variables were exhausted.

Our second goal was to assess whether changes in mobility were independently related to neighborhood percent positivity. Specifically, we hypothesized that neighborhood SES may influence neighborhood percent positivity through reduced ability to decrease mobility during the pandemic. To assess this, we first included a ‘total effect’ model with proportion uninsured as the explanatory variable of interest and adjusting for other SES variables as sources of confounding. Next, we added change in mobility to this model and compared (a) the change in the magnitude of the regression parameter estimate for uninsured proportion between the two models; and (b) assessed the magnitude and precision of the regression parameter estimate for change in mobility, adjusting for the SES variables. All models used linear regression with generalized estimating equations to account for within-ZIP correlation.

We report the results of the analyses at four time points during the study period, April 1, April 10, April 20 and April 30, using data available through the given day. For April 1 time point, as daily data are not publicly available before this date, the outcome was calculated from cumulative cases and tests for the month of March.

## RESULTS

277520 COVID-19 tests were performed across NYC during the month of April, and 124135 (44.7%) were confirmed positive, with a median (IQR) ZIP code-level positivity of 43.6% (38%-48.1%). Across all ZIP codes, the median (IQR) proportion of 18-64 year olds without health insurance was 13% (9-17%), median (IQR) household income was $63.1 ($47.2-$87.6) thousand (in 2016 US dollars), 25% of the median ZIP code population lived in households of 4 or more (IQR: 15.1% - 30.7%), about half of the population identified themselves as being white (25%- 68%), 9.7% (6.5-15) of those who relied on public transportation to commute to work used buses for part of their commute, and 11.8% (9.8%-14.4%) of the population was found to be elderly. Figure 1 shows distribution of these neighborhood SES characteristics by ZIP code.

**Figure 1.**
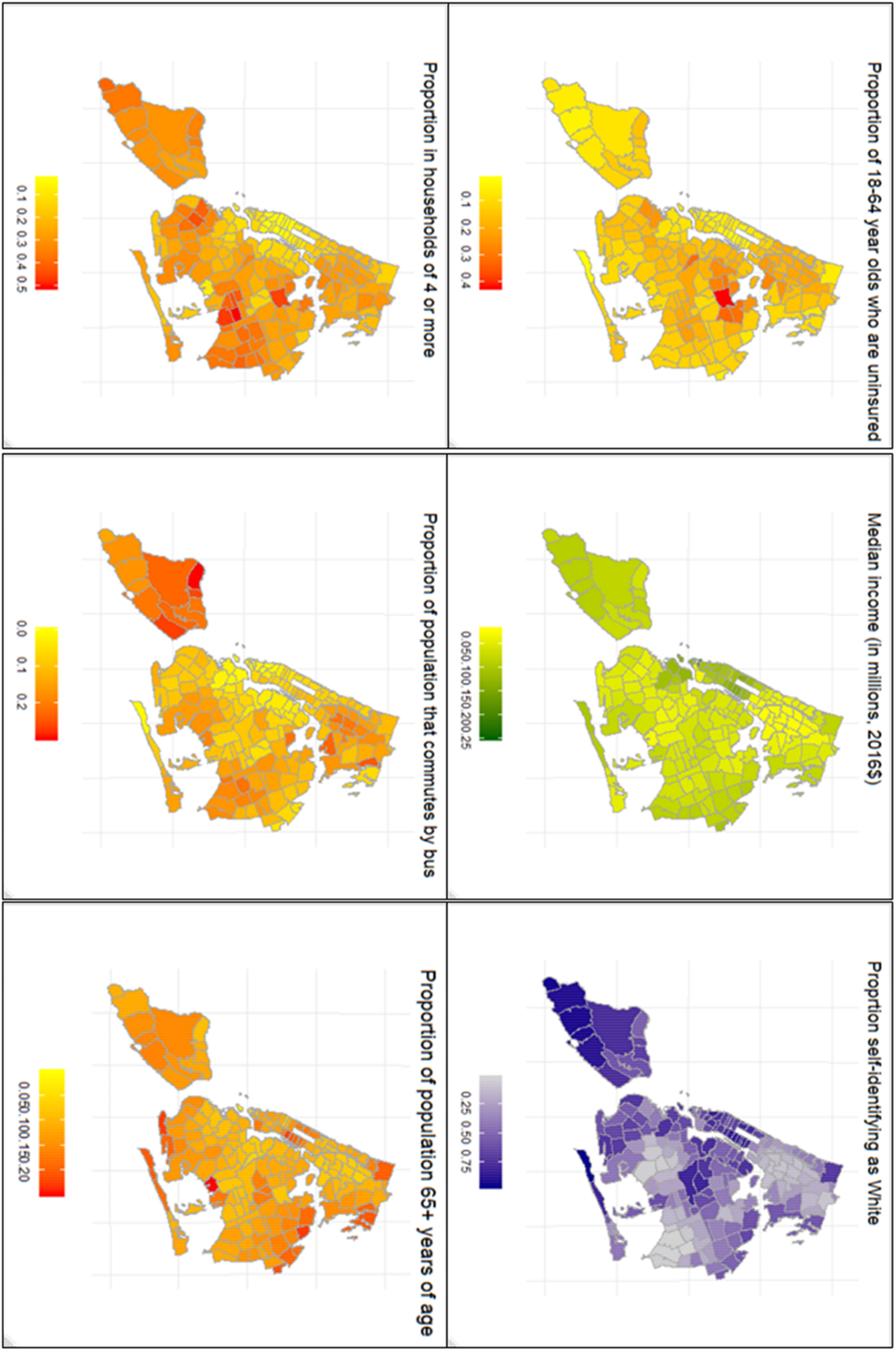
Maps of six explanatory variables used in this study as measured of SES characteristics of ZIP codes.

Figure 2 presents the proportional change in mobility over time by ZIP code. Median daily mobility remained stable prior to the official closures mandated by NYS on PAUSE going into effect on March 22, 2020. Between March 1-March 21, compared to pre-pandemic baseline, a median change in mobility of −8% (IQR: −25% to +14%) was observed. Following the mandated closures, a pronounced decrease in median mobility was observed, with the largest median decrease in mobility of −64% (IQR: −69% to −58%) on April 12^th^. Between March 21-April 19 median mobility remained relatively stable, but a trend toward increasing median mobility was noted from April 21-30. There is also a discernable weekly pattern in the mobility, with a lower mobility observed during weekends than on weekdays.

**Figure 2.**
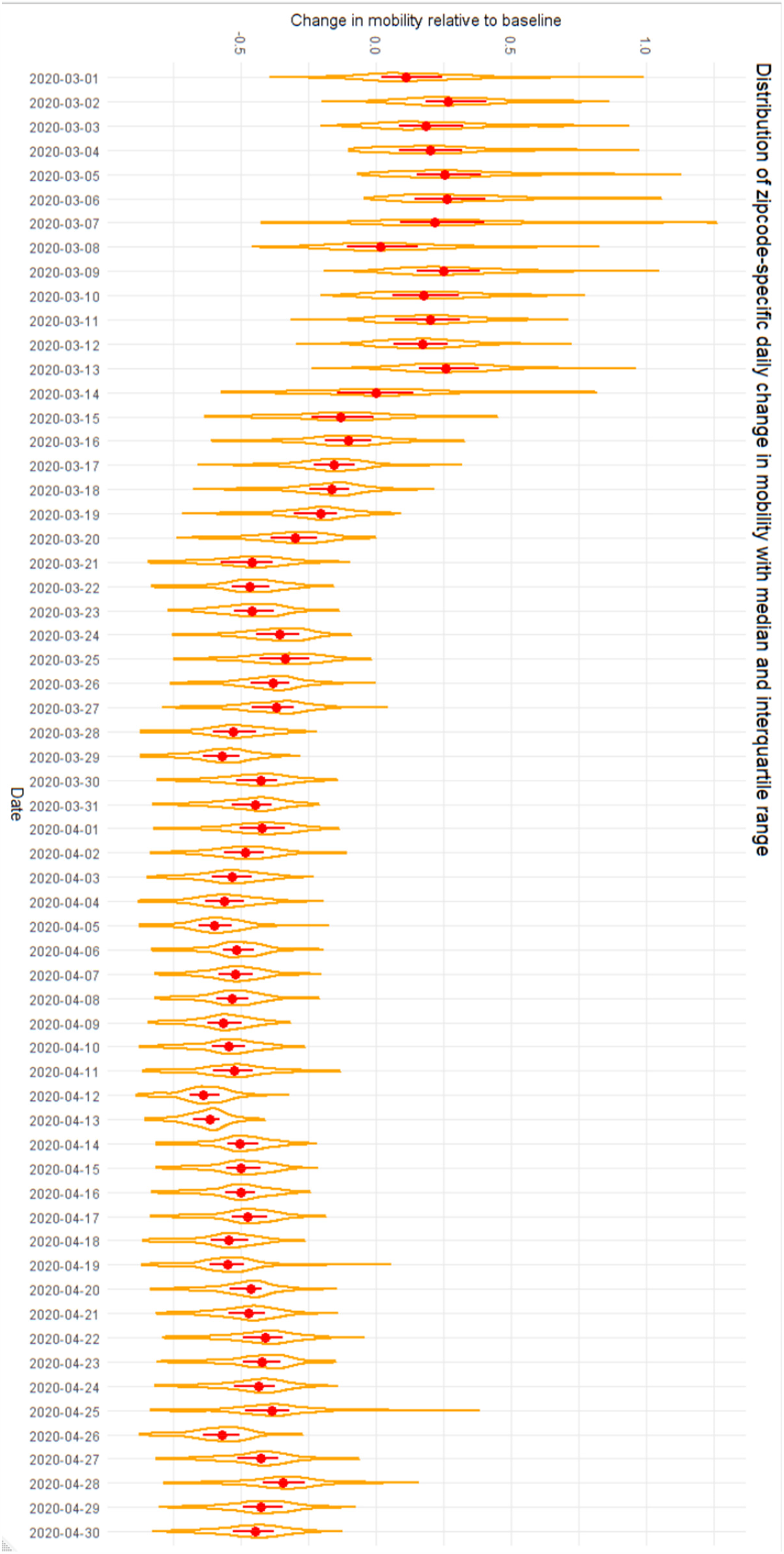
Change in mobility by date. The bounded area represents the distribution of change in mobility across ZIP codes. The median and interquartile range are shown in red.

Table 1 presents the results of the model explaining variability in positivity by ZIP code. In univariate analysis, at the first analysis point of April 1, 41% of the total variability by ZIP code in COVID-19 positivity was explained by a linear relationship with the proportion of the ZIP code living in a household with 4 or more individuals (R^2^ = 41%). R^2^ for the remaining explanatory variables assessed are: proportion of 18-64 adults who are uninsured (38%), proportion of population self-identifying as white (34%), median income (32%), change in mobility (19%), proportion using bus for commute (13%) and proportion elderly (3%). Sequentially including variables in our prediction model in order of decreasing R^2^ resulted in a model with three SES variables that explained 56% of the total variability in COVID-19 positivity: proportion living in households with 4 or more individuals, proportion of adults who were uninsured, and proportion identifying as white. Further addition of variables resulted in marginal changes.

**Table 1.**
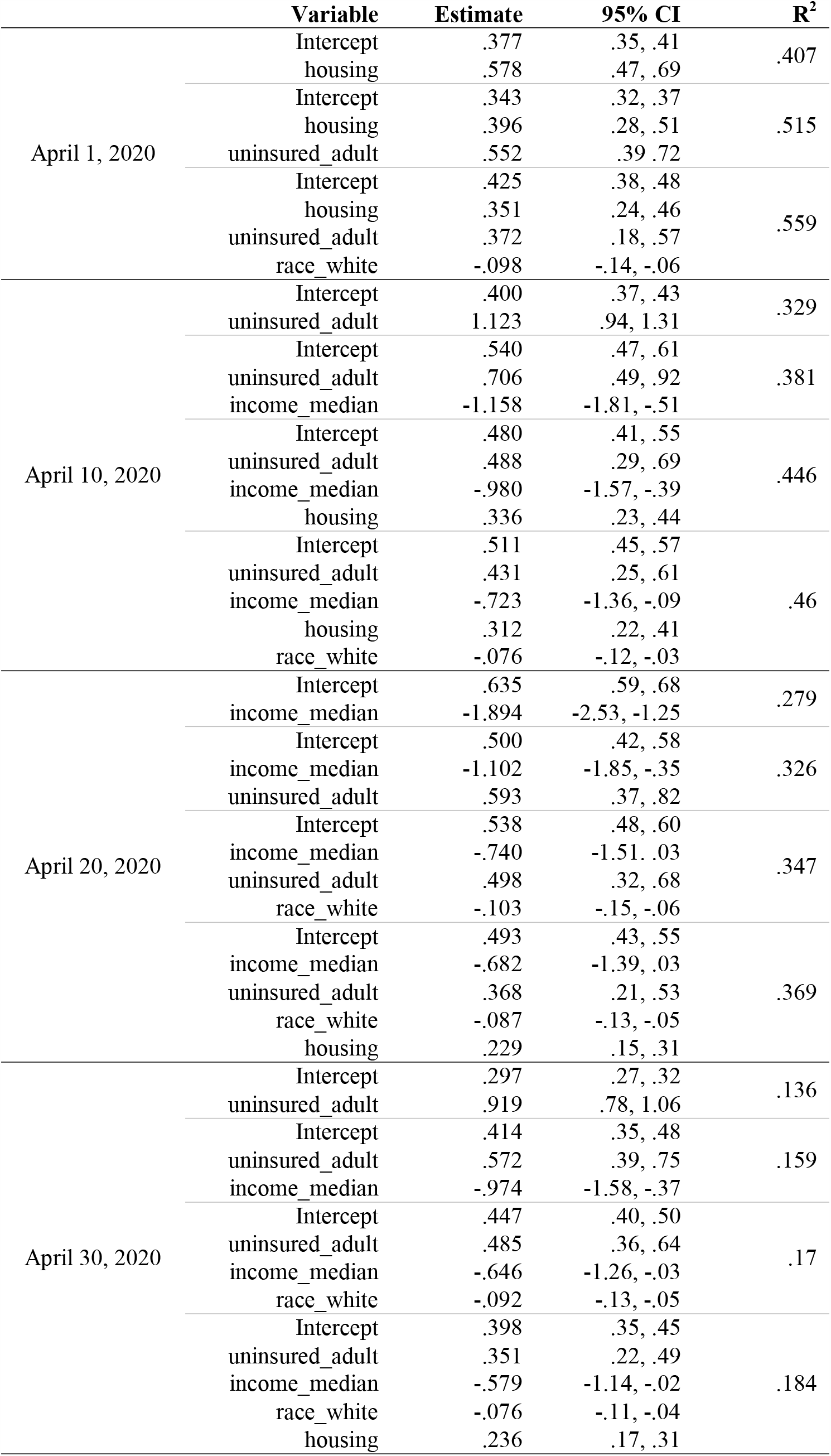
Results of univariate analysis at April 1, April 10, April 20, April 30

In analyses conducted at later time points, median income emerged as the variable explaining the most or second-most variability in COVID-19 positivity through univariate analyses. The other variables explaining substantial variability on April 1 remained. The explanatory power of these variables, assessed together, decreases over time from a model R^2^ from 56% (April 1) to a model R^2^ of 18% (April 30).

Following the observation that the proportion of uninsured adults is the most or second-most important variable in univariate analysis at all four time points, we examined the independent relationship between the proportion uninsured and percent positivity, adjusting for variables thought to act as confounders of the proportion uninsured → COVID-19 positivity relationship (Table 2). Adjusting for proportion over the age of 65 years, proportion whose work commute included bus travel, proportion living in households with 4 or more individuals, median household income, and proportion self-identifying as white, for every 10% increase in proportion of uninsured residents, percent positivity through April 1 increased by 3.2% (95% CI: 0.5%, 5.9%). The magnitude of this adjusted relationship between proportion uninsured and COVID-19 positivity was relatively stable across time points assessed -- 4.6% (2.5%, 6.8%) on April 10, 3.8% (2.1%, 5.6%) on April 20, and 3.9% (2.5%, 5.3%) on April 30.

**Table 2.**
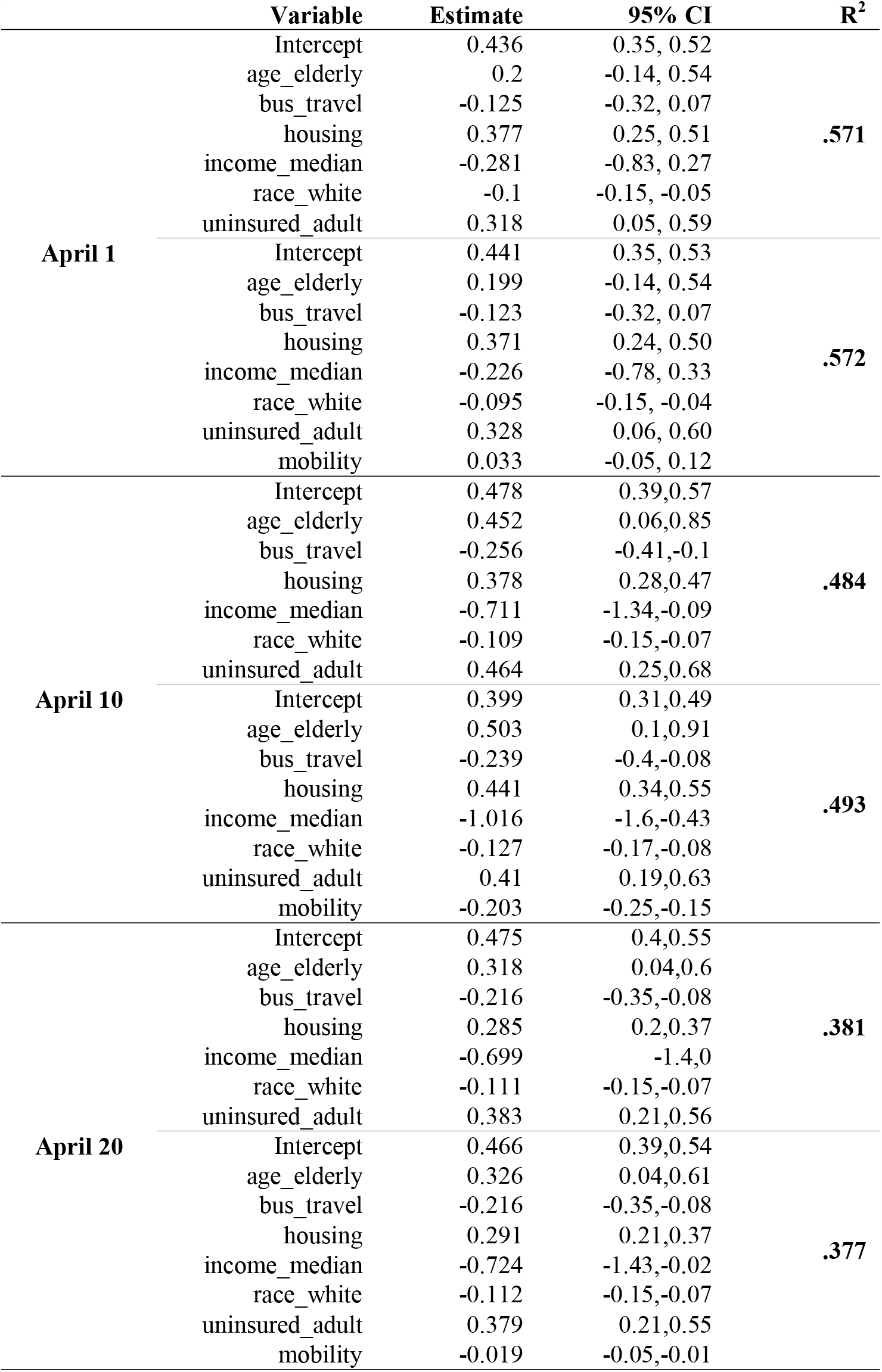
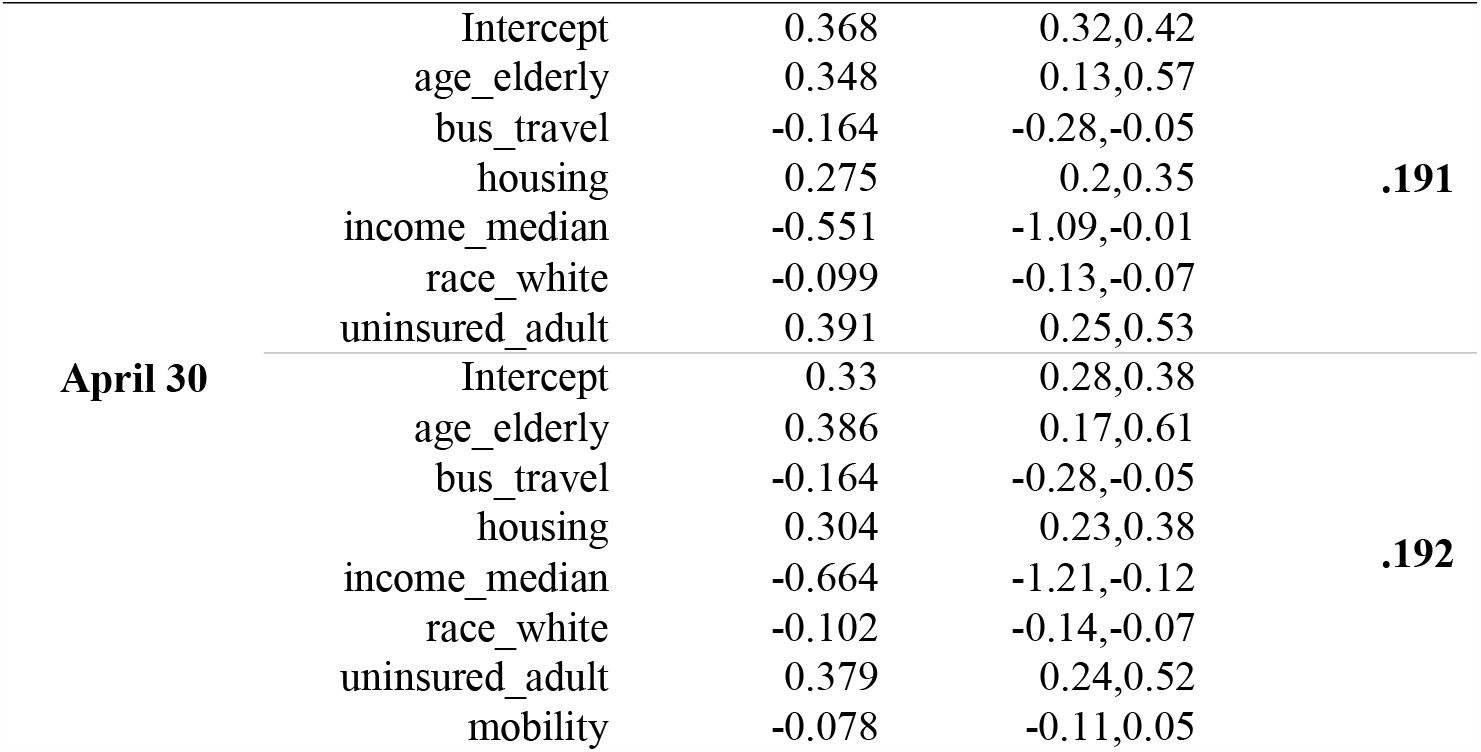
Results of regression analysis with Generalized Estimating Equations at April 1, April 10, April 20, April 30

In analyses of the possibility that changes in mobility may partially explain the observed SES→COVID-19 positivity relationship, we compared the adjusted regression parameter estimate for proportion uninsured in models with and without measure of change in mobility. After additionally including mobility, the estimate for uninsured proportion was largely unchanged with the exception of the April 10 time point (percent change in regression parameter estimate for proportion of adults uninsured in models with and without change in mobility: April 1: 3% increase, April 10: 12% decrease, April 20: 1% decrease, April 30: 3% decrease) (Table 2).

Finally, we assessed whether change in mobility was independently associated with case positivity at four time points corresponding to the pre-peak phase of the epidemic (April 1), shortly after the peak (April 10), and during the reduction in daily cases (April 20 and April 30). In unadjusted models, on April 1, ZIP codes differing in change in mobility from baseline by 10% had mean case positivity proportions differ by 3.1% (95% CI: 2.2%, 4.4%), with the higher case positivity among ZIP codes reducing mobility less. After adjusting for covariates (Table 2), change in mobility on April 1 was no longer independently associated with case positivity. At later time points, smaller reductions in mobility from baseline were associated with lower case positivity on April 10 and 20 but not on April 30. For example, on April 10, ZIP codes similar on covariates but differing only by a 10% difference in reduction in mobility are expected to have case positivity proportions differ by 2.0%, with the higher case positivity among ZIP codes changing mobility more.

## DISCUSSION

Our findings indicate that the heterogeneous distribution in neighborhood level case positivity during the first two months (of the presumably first wave) of the COVID-19 pandemic in NYC largely followed underlying SES markers. We also found that spatial differences in change in mobility were independently related to case positivity during the middle of April—with a smaller reduction in mobility independently associated with reduced positivity rates — but not at the beginning and the end of the month. A likely explanation for this is increased COVID testing during April. At the beginning of April, routine testing for COVID was not widely available in NYC and likely differed by ZIP code in its availability. At the same time, change in mobility was the most dramatic during the early parts of April. However, as testing became more widely performed, case positivity decreased overall, leading to an overall inverse association between change in mobility and reduction on COVID positivity. Alternatively, this could be due to differences in how quickly ZIP codes adapted to shelter in place guidelines even if, by the end of April, the overall reduction in mobility was similar across ZIP codes. Analysis of change in mobility as a potential mediator of a SES→case positivity relationship yielded mixed results, with a 12% reduction in the independent relationship between proportion uninsured and case positivity on April 10 but minimal differences elsewhere.

Across all analyses there remained a strong relationship between markers of socioeconomic status and case positivity by ZIP code. These findings align with a host of literature on the disproportionate impact of acute events on lower SES communities (13-15) and highlight the need for more targeted interventions. Neighborhoods with a larger proportion of uninsured and hence with more limited access to healthcare were found to have higher case positivity rates. Consequently, planning for future waves of the COVID-19 pandemic, or future pandemics, would benefit from early and universally affordable access to testing.

Our analysis found that during the peak of the epidemic, and early on in the city’s response to the pandemic, differences in COVID-19 positivity by ZIP code could largely be explained by static markers of neighborhood socioeconomic status. Over half of the variability in positivity on April 1, for example, was explained by just three measures (the proportion in a ZIP code living in a house with 4 or more members, the proportion of adults uninsured, and the proportion self-identifying as white). Note that daily COVID-19 cases peaked on April 6 in NYC. As the epidemic progressed, these markers of socioeconomic status, while still independently associated with positivity, became less predictive. Taken together, this is consistent with two plausible mechanisms: (1) lower case positivity among higher SES ZIP codes may in part be explained by greater ease in being able to receive tests despite not being clinically warranted among individuals in higher-SES neighborhoods, and (2) higher actual numbers of infections among individuals in lower SES ZIP codes. While our outcome measure of interest (case positivity) cannot definitively separate out these mechanisms, both can be important drivers of the heterogeneous experience of the epidemic across New York City.

Similarly, the importance of household density in these models underscores what is known from studies on transmissibility of other respiratory infections, including influenza, that most infections occur among members of a household (or in workplaces) due to prolonged exposure to infected individuals. Dense housing also limits the ability of those known to be infected to self-isolate; providing infected individuals the option to isolate at a location different from home could prove beneficial in these cases.

Our analysis showed that mobility reduced quite rapidly across all ZIP codes, beginning before mandated restrictions from NYS on PAUSE. This strongly suggests that many city residents dramatically curtailed their activities. The rate and magnitude of reduction varied by neighborhood, but higher reductions in mobility were actually independently associated with higher case positivity on April 10 and April 20 but not on April 1 or April 30. These mixed findings are partially due to the correlation between time and overall city-wide case positivity; as testing became more prevalent NYC observed a large decrease in overall case positivity.

Our study has a number of limitations. First, this is an ecologic analysis and thus inference is limited to the ZIP code-level, not the individual level. ZIP code is not a perfect measure of neighborhood and can mask some important heterogeneity in both exposure and outcome measures in this study. Second, we are unable to adjust for differential changes in population density by ZIP code as individuals with the means to leave NYC during this time period were likely to be disproportionately those of higher SES (16). As such, our measure of mobility cannot distinguish between ZIP codes having fewer visitors who individually have not reduced their visitation frequency, from a ZIP code with a constant volume of visitors who on average reduced their visit frequency. Third, limitations in the availability of COVID-19 positivity by ZIP code at the beginning of the pandemic limits our ability to fully understand the relationship between ZIP code-level mobility, SES, and positivity. We observed that the majority of the reduction in mobility occurred before public positivity data were available. Availability of ZIP level COVID-19 positivity in March 2020 would greatly strengthen our understanding. Finally, COVID-19 positivity is an imprecise outcome measure as it is heavily influenced both by the overall COVID-19 prevalence in a given ZIP code and access to diagnostic tests. Furthermore, the daily case and test counts were calculated as the difference between two successive *cumulative* case and test counts and hence it is assumed that the increments in cumulative counts are new case/tests and not revisions to counts from previous days.

Our study also has several important strengths. First, it uses a novel approach to measuring mobility that does not focus on distance travelled or average distance from presumed home location of a cell phone, but instead focuses on physical check-ins at POIs within an individual’s presumed neighborhood. As such, this metric is conceptually different measure of ‘mobility’ than other tracking measures, and one now being used by the CDC(11). Second, our simple model effectively explains a high amount of the total variability in COVID-19 positivity: on April 1 a model including only three variables (proportion living in a household with 4 or more individuals, proportion of 18-64 year olds without health insurance, proportion identifying as white) explained 56% of the total variability in COVID-19 positivity. While the explanatory power of the model decreased over the time span of the first wave of this pandemic, it was still able to explain substantial variability. Third, our model was robust to changes in the assumed lag time between exposure and outcome: models changing the lag from 7 days to 5 and 10 days yielded no substantial differences. Finally, the finding that change in mobility was independently associated with case positivity during the middle section of the epidemic response, but not at the beginning or the end of the time period of interest, suggests that more structural issues dominate the heterogeneous impact of COVID-19 in NYC. Further analyses as NYC begins re-opening will be useful to assess the degree to which returns to pre-COVID mobility impact the distribution of future waves of this pandemic.

## Conclusions

Dramatic reductions on mobility following ramp-up of governmental interventions strongly suggests that the NYS on PAUSE worked as intended, although more recent data indicative of increases in mobility suggest that there may be a point at which individuals begin relaxing their adherence to these interventions. Evidence from the first wave at the epicenter of the US COVID-19 pandemic strongly suggests that COVID-19 incidence is unequally distributed, largely by socioeconomic status. Intervention efforts should target communities most in need.

## Data Availability

Data is publicly available data obtained from the following sources:
https://github.com/nychealth/coronavirus-data
https://www.census.gov/acs/www/data/data-tables-and-tools/data-profiles/2016/
In addition, data on mobility was obtained from the following source and is available for purchase:
https://www.safegraph.com/

## Acknowledgment

We thank SafeGraph for sharing mobility information. SafeGraph is a data company that aggregates anonymized location data from numerous applications in order to provide insights about physical places. To enhance privacy, it excludes census block group information if fewer than five devices visited an establishment in a month from a given census block group.

